# COVID-19 preventive measures in Rohingya refugee camps: An assessment of knowledge, attitude and practice toward COVID-19

**DOI:** 10.1101/2023.02.21.23286227

**Authors:** Charls Erik Halder, Md Abeed Hasan, Yusuf Mohamed, Marsela Nyawara, James Charles Okello, Md Nahid Mizan, Md Abu Sayum, Ahmed Hossain

## Abstract

**Background:** Although many studies were conducted on COVID-19 knowledge, attitude, and practice among the general population in many countries, very little is known about refugees, particularly Rohingya refugees in Cox’s Bazar. A vast array of Risk Communication and Community Engagement (RCCE) interventions were implemented in Cox’s Bazar with the intent of reducing disease transmission by empowering the community to adopt public health measures.

**Objectives:** The study aimed to evaluate the current state of knowledge, attitude, and practice among Rohingya refugees as a result of RCCE initiatives.

**Materials and Methods:** A cross-sectional study was conducted with 500 Rohingya individuals. Participants in the study were Rohingya refugees residing in five randomly selected camps where IOM health was operating. Using a structured questionnaire, skilled community health workers surveyed the Rohingya population. In addition to the survey on knowledge, attitude, and practice, the study gathered information on the perspectives and relevance of sociodemographic factors that influence KAP.

**Results:** The study findings indicate that the mean scores for knowledge, attitude, and practice were 9.93 (out of 14), 7.55 (out of 11), and 2.71 (out of 7) respectively. Association was found between knowledge and practice level and age group – elderly age group (>/= 61 years) had less level of knowledge (AOR 0.42, P value= 0.05) and the late mid age group (46 – 60 years) had better practice level (AOR 2.67, P value <0.001). A significant association was also found between good knowledge level and medium family size (5 – 6 members) (P value= 0.02).

**Conclusions:** The study reveals that the Cox’s Bazar Rohingya refugee community has a low knowledge and attitude score about COVID-19 prevention measures. Especially the KAP scores were found significantly low in elderly population. Despite RCCE interventions, the practice level of these measures exhibited a considerably low score. Poor implementation of preventive measures must be identified and remedied, involving the community in future outbreaks of a similar nature.

## Introduction

Approximately 883,600 Rohingya refugees are residing in 34 overcrowded refugee camps in Cox’s Bazar following their mass displacement from Myanmar in 2017 [1], preceded in kind by decades of influxes spurned by systematic discrimination and targeted violence [2]. The refugees are living in overcrowded bamboo-made settlements at hilly slopes and basins with limited access to livelihood and basic entitlements and are highly vulnerable to natural and man-made disasters and disease outbreaks [3]. Crowded living conditions, lack of good WASH facilities and practices, and heavy monsoon in the refugee camps and adjacent host communities increase their susceptibility to infectious diseases and often result in disease outbreaks [4]. Since 2017, several outbreaks or upsurge of infectious diseases, like, diphtheria, measles, AWD/cholera and dengue were reported in the Rohingya camps [4]

Superimposed on the existing vulnerability to disease outbreaks, COVID-19 appeared as a new threat to this population. COVID-19 is a highly contagious emerging disease caused by Severe Acute Respiratory Syndrome Coronavirus 2 (SARS-CoV-2). The disease was first learnt in December 2019 following a report of a cluster of viral pneumonia cases in Wuhan [5]. Since then, the disease continued to spread around the world, with more than 500 million confirmed cases and six million deaths reported across around 200 countries [6]. On March 11, 2020. COVID-19 outbreak was declared by World Health Organization (WHO) as a global pandemic [7]. The first COVID-19 positive case confirmed in Bangladesh was on March 08, 2020, and the first case from the Rohingya refugee camps was reported on May 2020 [8, 9]. As of 24 April 2022, 5,922 COVID-19 confirmed cases (out of 99,049 tests) and 42 deaths were reported from the refugee camps [10].

Child health is at risk among Rohingya refugees due to inadequate child protection measures [11, 12]. The high prevalence of mental health issues among Rohingya refugees further complicates efforts to make them aware of the danger posed by the virus [13]. Public health guidance for prevention and control of COVID-19 included maintaining distancing of one meter from others, wearing a mask, cleaning hands, covering cough and sneezes, getting vaccinated, staying home when sick and seeking medical care when required [14, 15]. Effective implementation of those measures relies largely on what people know about these, how they think or believe on these and how they do or experience these, which in summary interpreted as knowledge, attitude, and practice (KAP) [16]. Before vaccine was widely available for the population, public health and social measures remained as the most important tool for interrupting the disease transmission [17]. Hence, risk communication and community engagement (RCCE) were one of the major pillars in COVID-19 response strategy [17]. IOM implemented a vast array of RCCE interventions in Cox’s Bazar with the intent of reducing disease transmission by empowering the community to adopt public health measures. The interventions included household visits and community meetings by community health workers, dissemination of audio-visual clips, publication of printed materials, social advocacy through social leaders and community groups, go and see visits to the service sites. Most messages and contents of the communication was based on materials developed by CwC working group.

Although many studies have been conducted on the knowledge, attitude, and practice of COVID-19 preventive measures among the general population in various countries, very little is known about the knowledge, attitude, and practice of refugees, particularly Rohingya refugees in Cox’s Bazar. Jubayer et. Al (2022) carried out a KAP survey COVID-19 among the Rohingya refugees at the very beginning of the outbreak [18]. However, the current status of the knowledge, attitude and practice is not known after undertaking the RCCE interventions. Different forums and reports have highlighted the issue of noncompliance among the population with COVID-19 measures; however, there is no evidence as to what extent the public health measures are not accepted or practiced by the community. The findings of the study could support the development of a robust strategy on risk communication and community engagement for the ongoing pandemic as well as future outbreaks of infectious diseases.

## Materials and Methods

### Study design and participants

The study was carried out in the refugee camps of Cox’s Bazar, where the International Organization for Migration (IOM) had implemented community health programs. The population of Rohingya community in Cox’s Bazar is around 883,600, with a male to female ratio of 45:55 [1]. Notably, a large portion of the population resides in makeshift shelters. It was a cross-sectional study. Rohingya refugees in the selected camps who were 18 years or above were the study participant. The inclusion criteria for this study were that participants must reside inside the camps, be 18 years or older, consist of both male and female participants who give their consent, and must be able to understand the questioner, while the exclusion criteria were that participants cannot belong to the same family. A survey was employed among 500 Rohingya individuals selected through systemic random sampling. 5 camps of IOM health operation were selected randomly. In each camp, 5 sub-blocks were randomly selected and at each sub-block, 20 households were selected through systemic random sampling. At each camp 5 CHWs surveyed 100 beneficiaries of different ages from the selected households; from each household, a beneficiary was selected using an age-sex table prepared based on camp demographic data.

### Sample size calculation

Since it was a cross-sectional study, the following formula used for sample size collection: 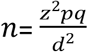

The equation represents a sample size calculation for our study. The variable “q” is calculated as 1-p, where “p” is the population proportion, which is assumed to be 50%. The variable “z” represents the confidence level of interest, which is set at 1.96 for a 95% confidence level. The variable “d” represents the degree of accuracy required, which is set at 0.05 level for the expected sample size. Using these values, the sample size “n” is calculated using the equation (z^2*p* q) / d^2, resulting in a sample size of approximately 384, rounded up to 500 to account for the risk of a 25% non-response rate. This sample size will be used to ensure that the study results have a high level of accuracy and confidence.

### Data collection instrument and data collection

The CHWs surveyed each participant using a questionnaire/checklist [Table 2] and by observing the practices in their day-to-day life. All data were entered into the kobo toolbox against the questionnaire by the CHWs. The questionnaire was validated by piloting among a small sample of participants to ensure that it is clear, relevant, and produces consistent and accurate data. The questionnaire comprised 14 questions to assess the respondents’ knowledge. Eleven and seven questions were employed, respectively, to evaluate the attitudes and practices of respondents regarding COVID-19. Respondents with a score of 9 or higher were deemed to have “good knowledge” of COVID-19, and an aggregate score was computed (range 0–14). Respondents with a score of 8 or higher were deemed to have a “positive attitude.” Measurements of cautious behavioral practices were based on the following two categories: (1) preventative measures (such as wearing masks and exercising hand hygiene) and (2) social isolation (i.e., avoiding crowded places, family staying at home). Using a two-point Likert scale (0 = No, 1 = Yes), CHWs monitored respondents’ actions during the survey period. Respondents with a score of 6 or higher were deemed to have “good practice” (Table 1a). The questionnaire/checklist was translated into the Bengali language.

**TABLE 1A:** METHOD OF CALCULATING KAP SCORE

| Variables | Number of Questions | Score to Answers | Level of Variables |
| --- | --- | --- | --- |
| Knowledge | 14 | 1= Yes/Correct<br>0= No/Don't know | Poor: 0-4 (0-33) |
|  |  |  | Moderate: 5-8 (34-67) |
|  |  |  | Good: 9-14 (68-100) |
| Attitude | 11 | 1= Yes/Correct<br>0= No/Don't know | Poor: 0-3 (0-33) |
|  |  |  | Moderate: 4-7 (34-67) |
|  |  |  | Good: 8-11 (68-100) |
| Practice | 7 | 1= Good practice<br>0= Poor practice | Poor: 0-3 (0-33) |
|  |  |  | Moderate: 4-5 (34-67) |
|  |  |  | Good: 6-7 (68-100) |

### Data analysis

To assess the level of knowledge, attitude and practice of COVID-19 preventive measures descriptive analyses of socio-demographic and exposure factors descriptive statistics were measured for each variable by using a similar scale developed by Zhong et al., the respondents’ knowledge, attitudes, and practices with COVID-19 were evaluated [16]. All responders were asked to respond “Yes” or “No”. Scores were determined by awarding one point for each appropriate answer, with higher scores signifying a greater proficiency level. The bivariate relationship between the socio-demographic and outcome variables was assessed using the Pearson chi-square test and Fisher exact test. Multivariable logistic regression was used to evaluate the status and effectiveness of risk communication and community engagement approaches, inter-activeness, acceptability, and comprehensibility. The logit coefficient is calculated, as well as the 95% confidence interval. The level of statistical significance was set at 5%. All analyses are carried out using the STATA (v-16.0).

### Ethical consideration

The Institutional Review Board (IRB)/Ethical Review Committee (ERC) of North South University, Bangladesh, authorized the protocol for this study (2021/OR-NSU/IRB/0401). All respondents participated voluntarily. Before performing the formal interview, each participant provided written informed consent (mainly thumb imprints), and consent documents were stored separately until the conclusion of the study in a closed filing cabinet. The study adhered to the “no-harm” principle, and there was no legal risk associated with the involvement of the participants. Moreover, local rules and regulations were observed during interactions. Each stage of this investigation was conducted in accordance with the Helsinki Declaration (1964) and its most recent amendment (2013).

## Results

### LEVEL OF KNOWLEDGE, ATTITUDE AND PRACTICE

#### Sample Characteristics of the study population

The demographics of the survey respondents in Rohingya refugee camps in Cox’s Bazar are shown in Table 1b. A total of 500 people participated in this study, with 239 men (47.80%) and 261 women (52.20%) making up the majority. The participants in the study were on average 43.98 years old. The study participant’s age categories are distinguished by early, prime, mature, and elderly [17]. This age group is classified by comparing the life expectancies of the host and refugee communities, with the average life expectancy for the Rohingya being 67.36 years and the Bangladeshi being 72.87 years [18]. In the study population, 20.40% were between the ages of 18 and 30, 35.60% were between the ages of 31 and 45, 28.80% were between the ages of 46 and 60, and 15.20% were above the age of 61. 89.20% of the people in the survey were married, 1.80% were single, and 9% were divorced or widowed. Families with fewer than five members accounted for 38.20% of respondents, followed by 34% for families with fewer than seven but more than four members, 21.20% for families with fewer than nine but more than six members, and 6.60% for families with more than nine members. 17.60% of respondents had two children, 23.60% had five or more children, 17% had three children, and 12.60% had one child, which was the same as those who did not have a child. One or more family members under the age of ten years were reported by 63% of respondents, while 26% had family members above the age of 60.

**TABLE 1B:**
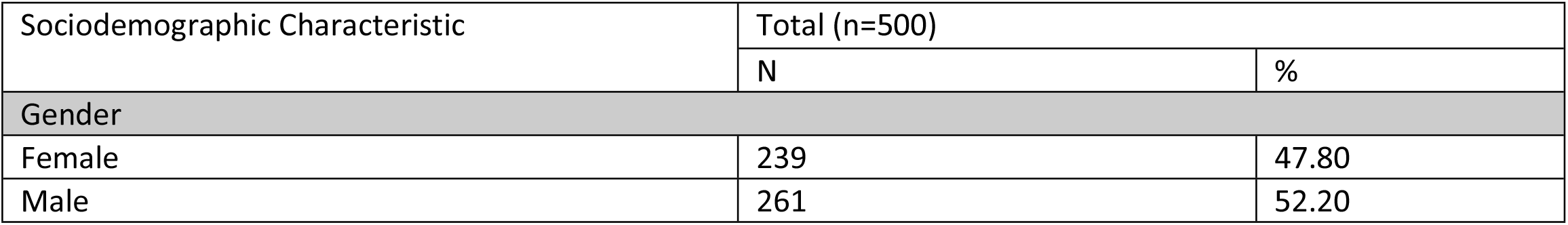
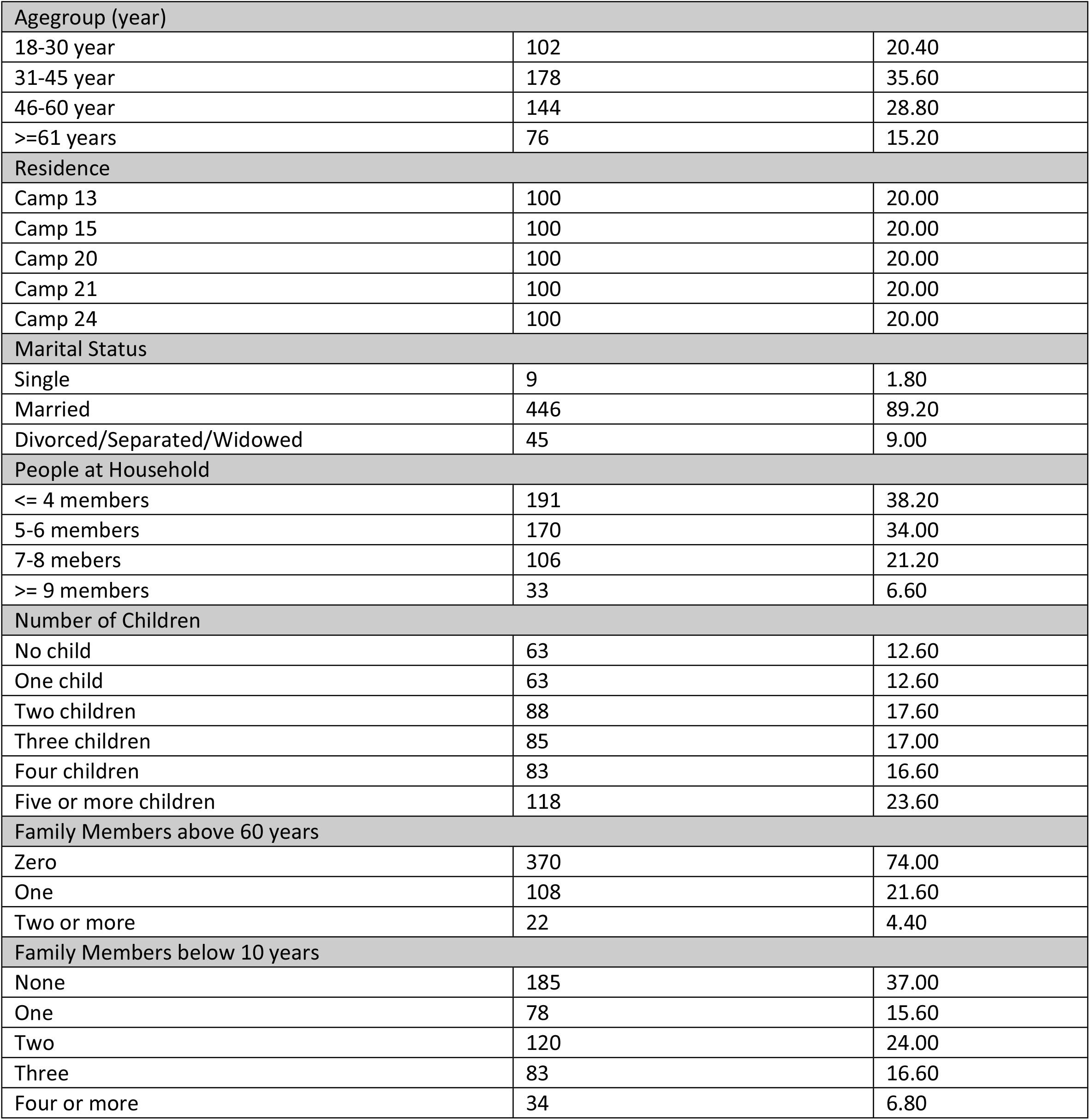
DESCRIPTIVE STATISTICS OF SURVEY RESPONDENTS (n= 500)

#### Level of knowledge, attitude and practice regarding COVID-19 preventive measures

Table 2 shows the response of the participants on their knowledge, attitude and practice regarding COVID-19 preventive measures. Majority of the participants had understanding of the different preventive measures for COVID-19. 100% of the participants heard about COVID-19 and more than 90% heard about the COVID-19 vaccine. More than 80% of the participants could explain some symptoms of COVID-19. Around three fourth of the participants understood how COVID-19 transmits, the necessity of consultation at the health facility for respiratory/COVID-19 symptoms, the requirement of wearing masks and cough etiquette. Two third of the participants knew that they should wear masks around other people and avoid crowded or closed spaces. Knowledge regarding complications and different severity of COVID-19 was relatively low, around half of the participants positively responded. Around 60% of the participants had knowledge regarding the necessity of providing samples for testing, isolation of positive cases at the isolation and treatment center, and quarantine of contacts of COVID-19.

**TABLE 2:**
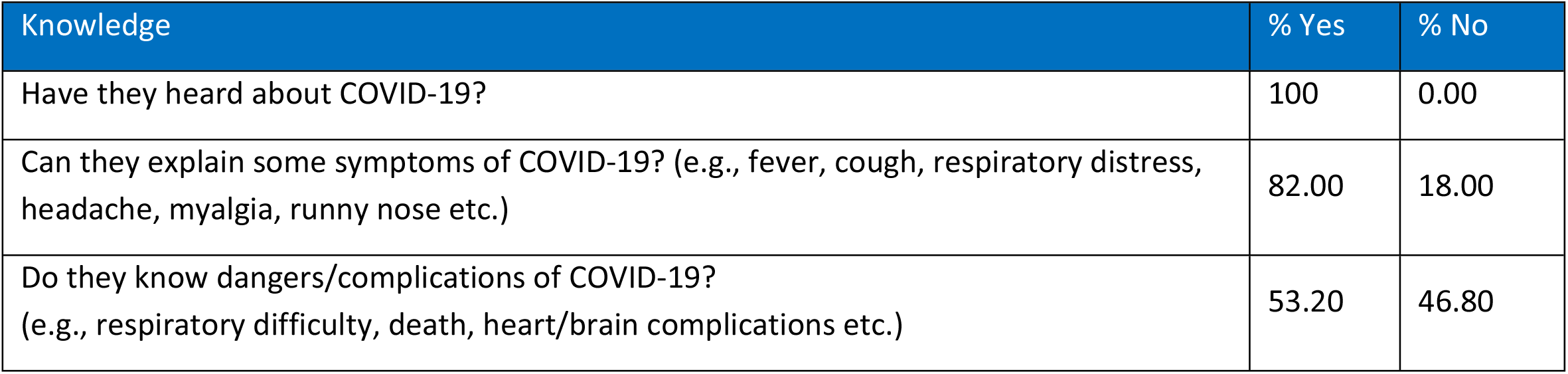
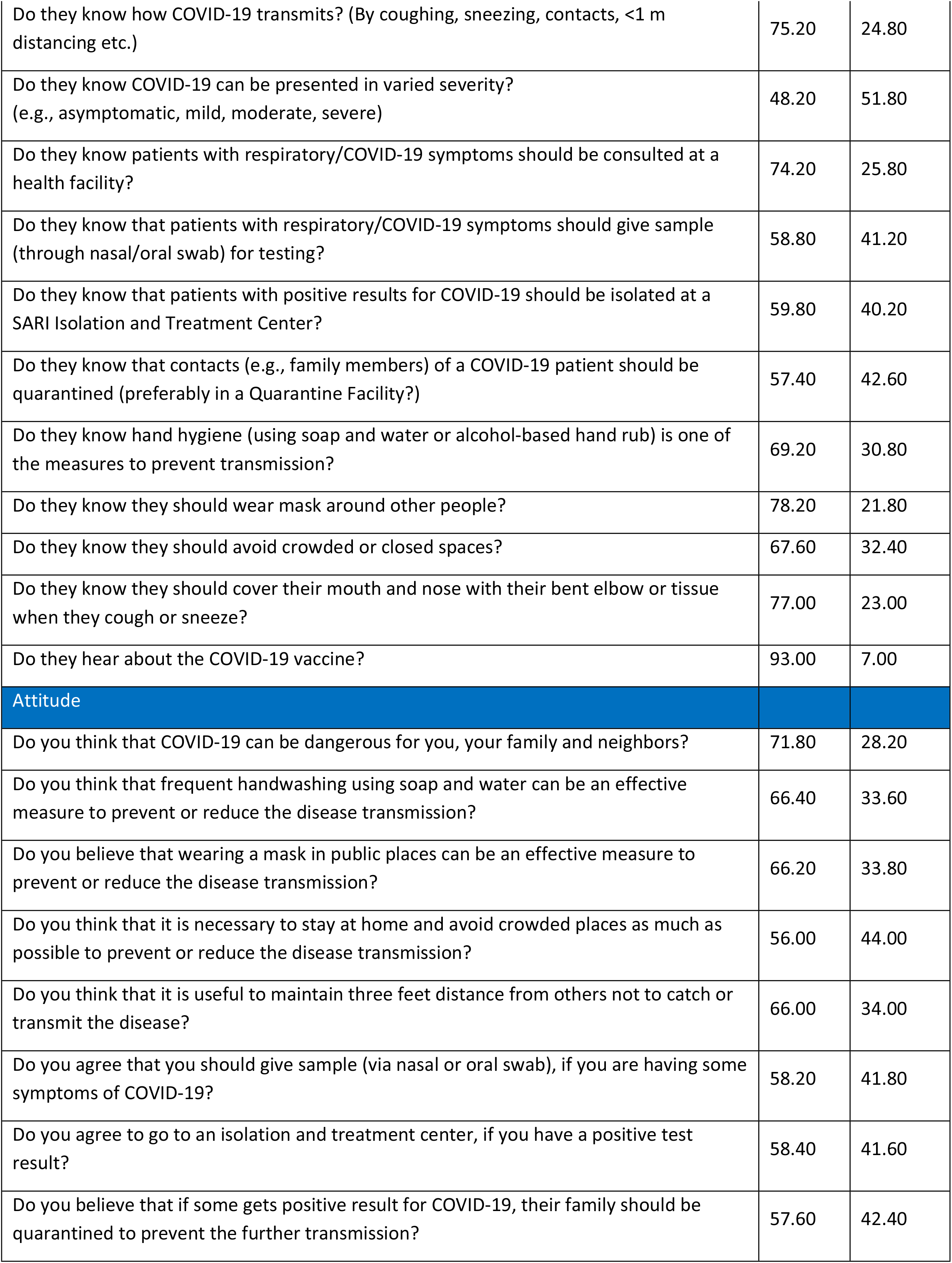
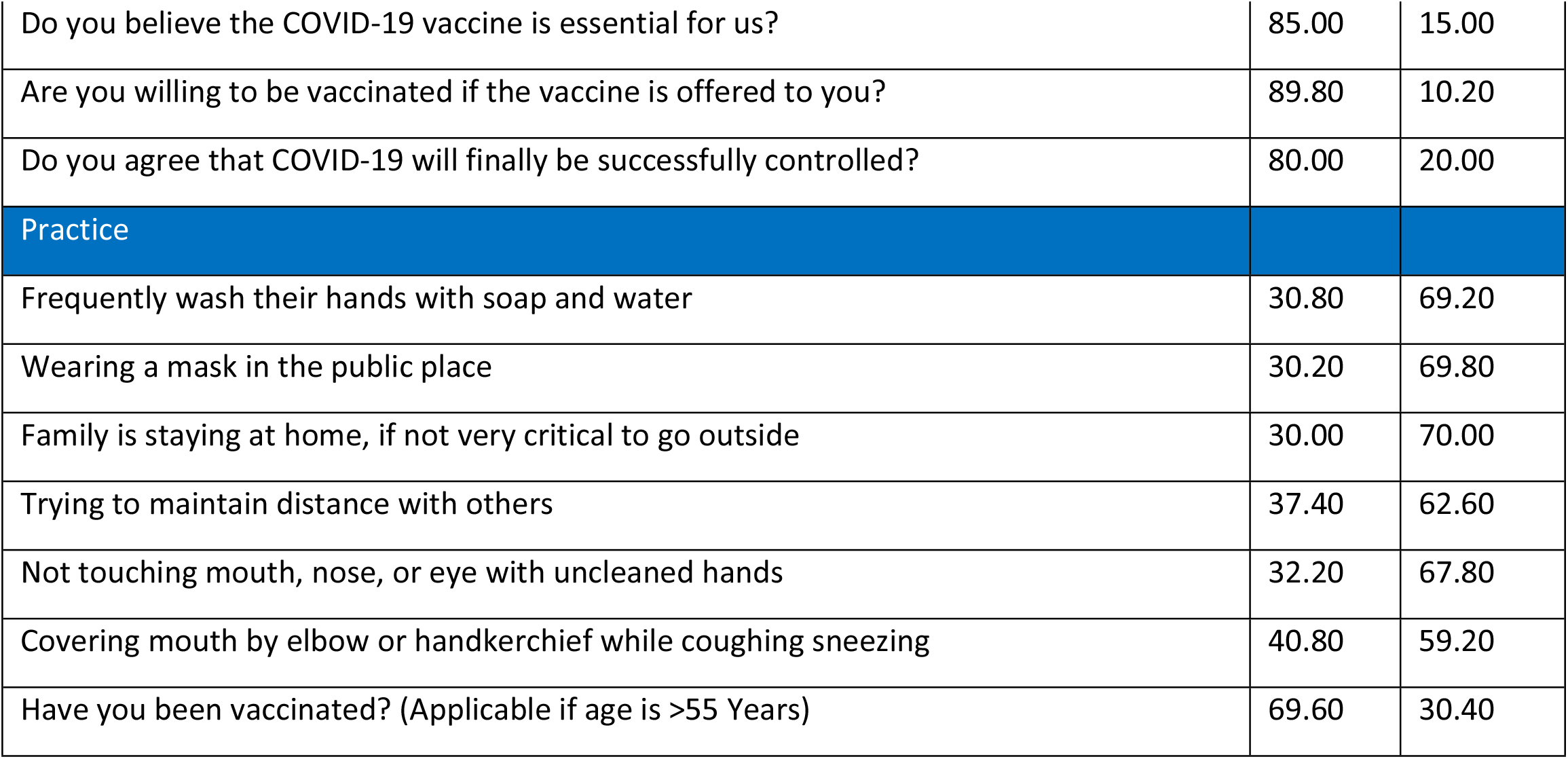
PARTICIPANTS’ KNOWLEDGE, ATTITUDE AND PRACTICES ON COVID-19 PREVENTIVE MEASURES

More than 70% of the participants perceived COVID-19 as dangerous for them and their families. Two-thirds of the participants had positive attitudes toward handwashing and wearing masks in public places and maintaining social distancing. The majority of the participants had positive attitudes toward the essentiality of COVID-19 vaccine (85%) and administering COVID-19 vaccine (89.8%). Below 60% of the participants agreed to avoid crowded places, giving sample if having symptoms, admission into an isolation and treatment center and quarantining the family members in case of a positive result. 80% of the participants agreed that COVID-19 could finally be controlled.

In comparison to knowledge and attitude, the practice of COVID-19 different preventive measures among the participants was found low. Below one-third of the participants frequently washed their hands, wore masks in public places, stayed home if not critical, and avoided touching their mouth, nose, or eye with uncleaned hands. Only 37% of participants tried to maintain distance from others and 40% practiced cough etiquette. However, around 70% of the participants, who were over 55 years, received a dose of COVID-19 vaccine.

Table 3 shows the results of the study in terms of level of knowledge, attitude, and practice. The findings suggest that many respondents (70.80%) had a good understanding of COVID-19, with only a minority (16.0%) had poor knowledge level. Only 22.20% of respondents had a good level and 37.02% had an average level of attitude toward maintaining COVID-19 preventive measures. One-fifth of the participants (22%) had a poor level of attitude. In terms of practice level, it was also found that only 7.00% of the respondents had a good level of preventive practices, while the majority of respondents (61.80%) had a poor level of practice. In summary, the majority of the community had a good level of knowledge or awareness on COVID-19 and an average to good level of attitude, however, a significantly low level of practice toward the preventive measures.

**TABLE 3:** KNOWLEDGE, ATTITUDE AND PRACTICE REGARDING COVID-19 PREVENTIVE MEASURES

| Variable | Overall Mean (+/- SD) | Poor | Average | Good |
| --- | --- | --- | --- | --- |
| Knowledge | 9.93 (2.95%) | 80 (16.00%) | 113 (22.60%) | 354 (70.80%) |
| Attitude | 7.55 (2.51%) | 111 (22.20%) | 186 (37.20%) | 111 (22.20%) |
| Practice | 2.71 (1.58%) | 309 (61.80%) | 201 (40.20%) | 35 (7.00%) |

Table-4 illustrates the association of the socio-demographic distribution of the respondents and knowledge, attitude, and practice regarding COVID-19 preventive measures. In terms of age, except for elderly people, almost all age groups had nearly the same percentage of responders (60∼70%) with good awareness of COVID-19 infection, prevention, and control strategies. Good attitude toward COVID-19 preventive measures was found similar among all age groups, between 38 – 44%. While the prevalence of the good practice of COVID-19 preventive measures was found low in all age groups (2 ∼ 11%), it is lower (3.92%) among the early working age group (18-30 year) and lowest (2.63%) among elderly (>/= 61 years). It was found that almost an identical proportion (∼62%) of the male and female participants have a good level of knowledge. In terms of the gender distribution of good level of practice and attitude, a bit lower prevalence was found among female participants in comparison to male.; 38.08% of the female participants had good attitude which is lower in comparison to the male (42.15%), and 6.28% of the female participants had good practice level which is lower in comparison to male (7.66%). We found that those who were separated/widowed (53.33%) had less understanding of COVID-19 than the single (77.78%) or married respondents (62.33%). Single respondents (66.67%) had a more positive attitude than married (39.91%) or separated/widowed respondents (37.78%). Positive attitudes toward preventive measures were found lower among married (39.91%) individuals and divorced/separated/widowed in comparison to singles (66.67%). Good practice level was also found lower among married (7.40%) and lowest among divorce/separated/widowed (2.22%) in comparison to singles (11.11%). We observed that 56.02% of respondents from small/nuclear families had a good level of knowledge, while 84.50% of respondents from very big families had a good level of knowledge. Good attitude level remains within 38 – 45% among respondents from different family sizes. However, good practice level was noted among 9.42% of respondents from small families and 3 – 4 % among large and very large families. Through bivariate analysis, the study found a significant association between family size and knowledge level (Fisher’s exact 0.02), however, no statistical association of knowledge, attitude and practice were found with gender and marital status.

**TABLE 4:**
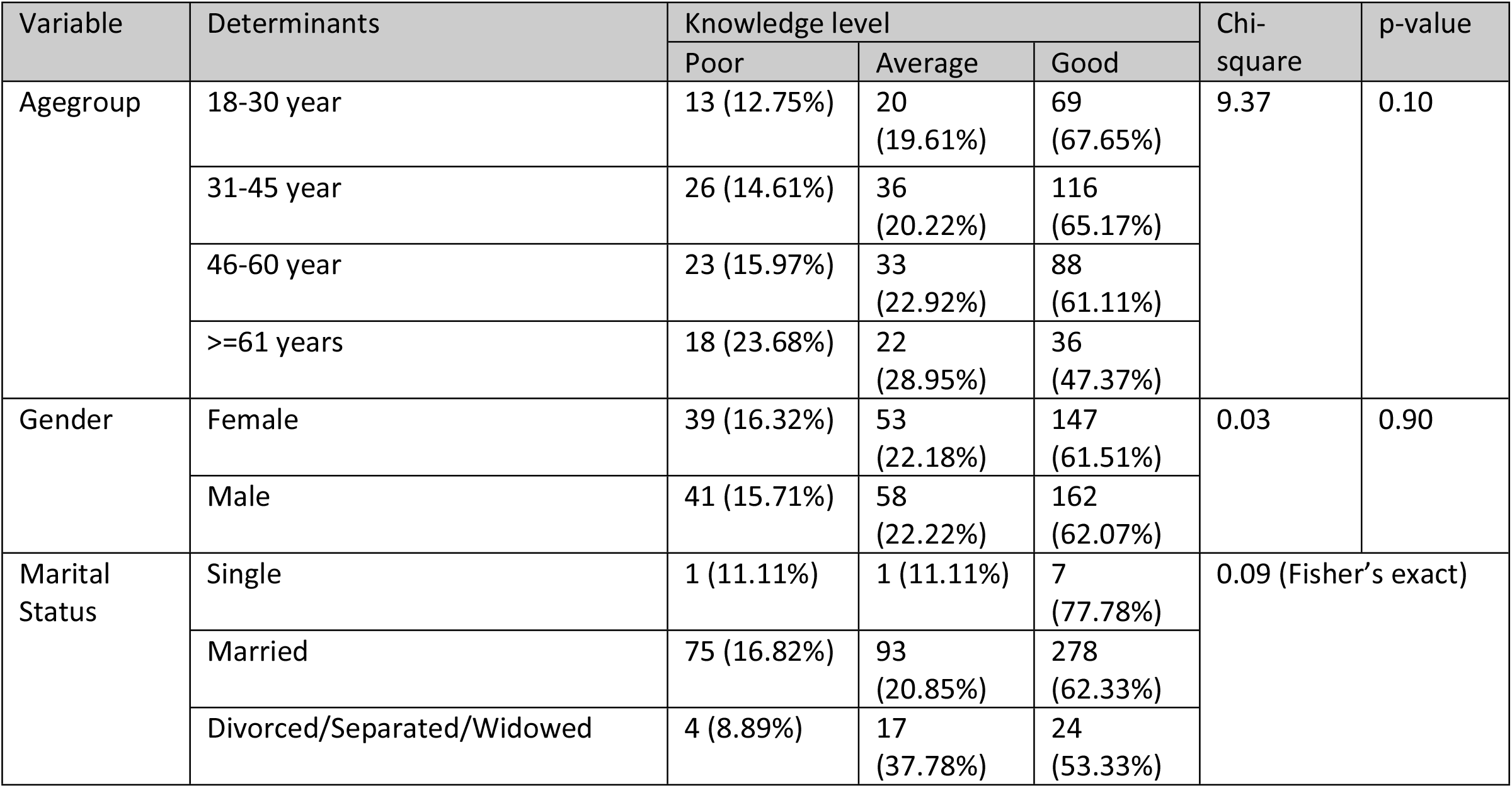
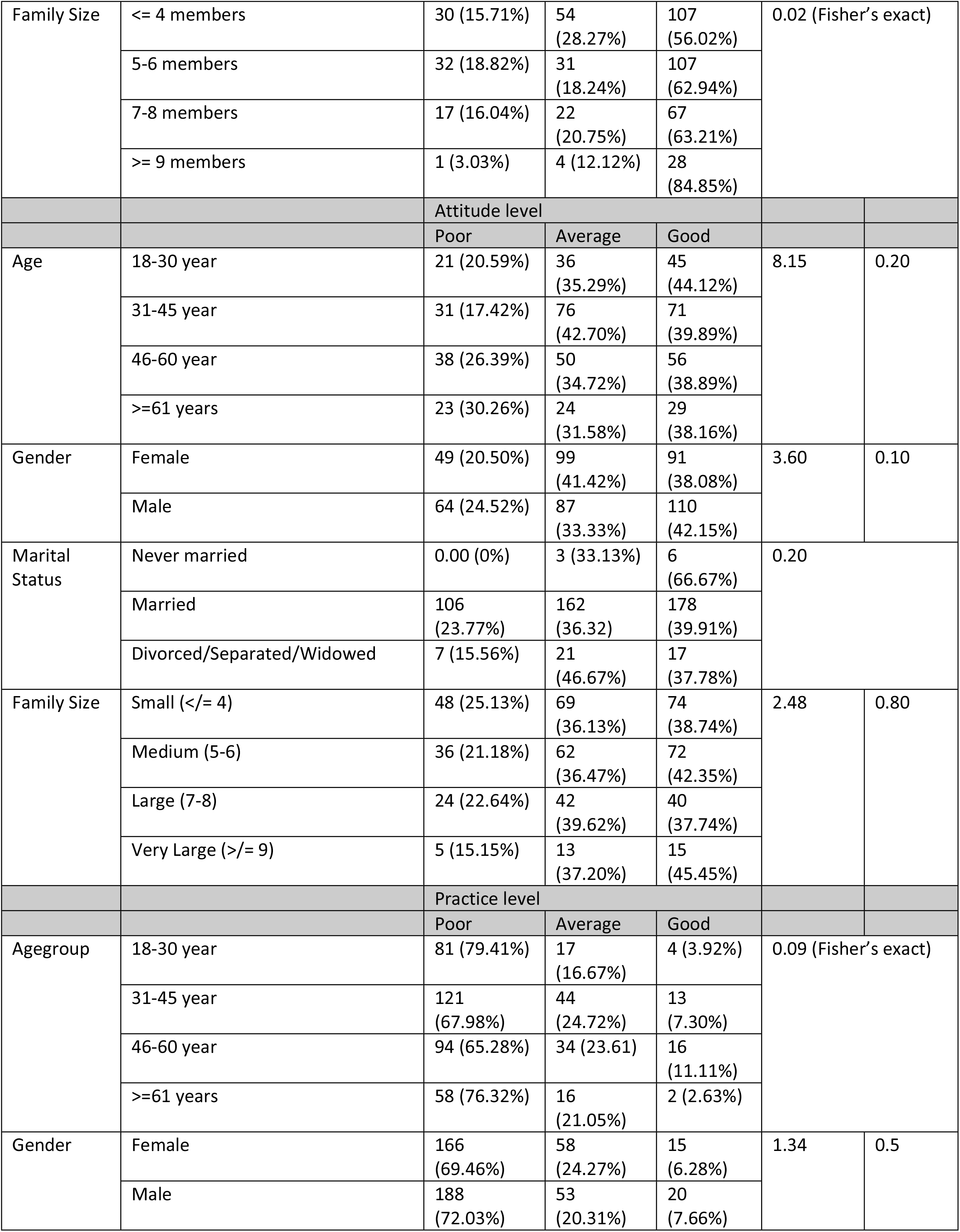
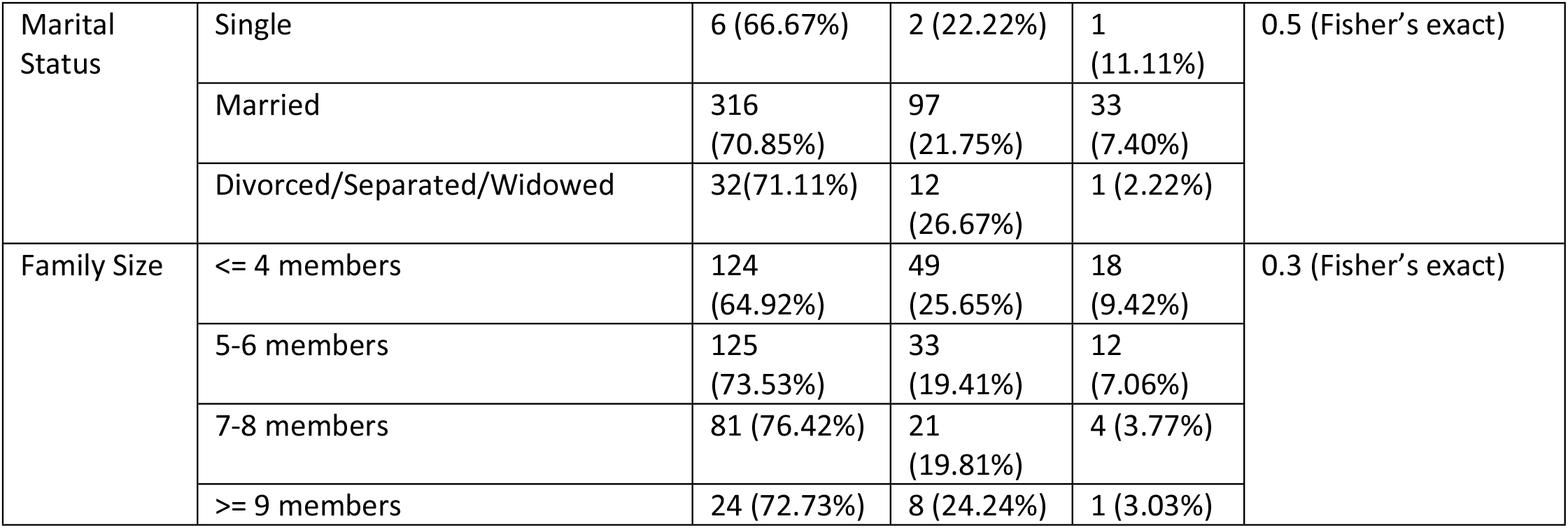
DISTRIBUTION OF RESPONDENTS AND ‘KNOWLEDGE, ATTITUDE AND PRACTICE’ SCORES OF COVID-19 ACROSS DEMOGRAPHICS IN FDMN CAMPS (N=500)

The scores of KAP were separated into three classes and placed as the dependent variable using the quartile as a cutoff point. The model is comprised of three continuous processes that aid in the explanation of human behavior residing in the camps. Table 5 illustrates in the multivariable logistic regression analyses, there is no statistical relevance with gender and marital status on KAP scale in terms of knowledge, attitude and practice regarding COVID-19 preventive measures. Association was found between knowledge and practice level and age group – elderly age group (>/= 61 years) had less level of knowledge (AOR 0.42, P value 0.05) and the late mid age group (46 – 60 years) had better practice level (AOR 2.67, P value 0.00). A significant association was also found between good knowledge level and medium family size (5 – 6 members) (P value, 0.02).

**TABLE 5:**
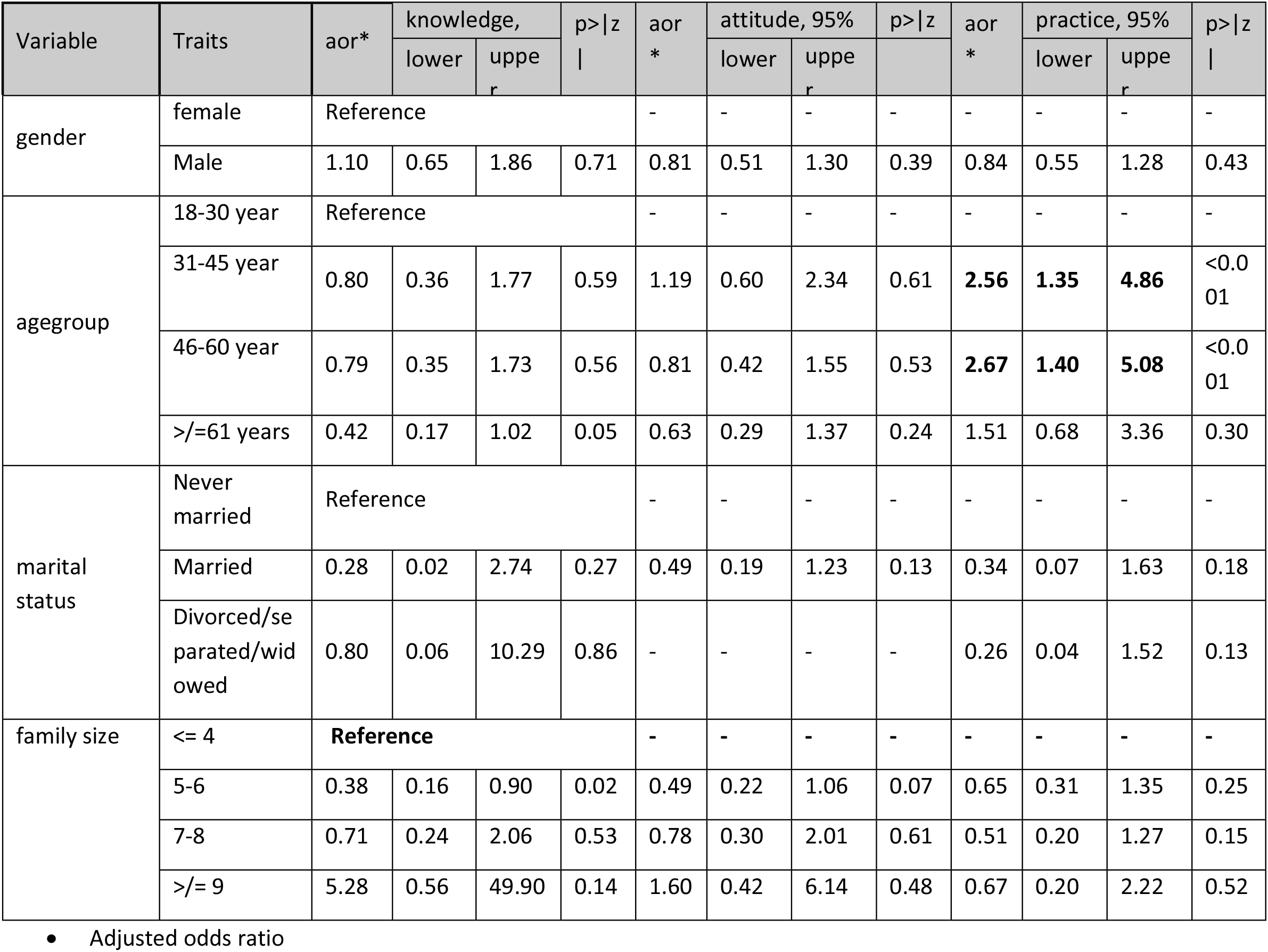
SIGNIFICANT FACTORS ASSOCIATED WITH KNOWLEDGE, ATTITUDE AND PRACTICE REGARDING COVID-19 PREVENTIVE MEASURES.

## Discussions

This study aimed to assess the level of knowledge, attitude, and practice concerning COVID-19, as well as the perceptions and importance of sociodemographic characteristics that influence KAP. With an unbiased distribution of gender, age, family size and marital status, this study aims to correlate with social determinants of health factors associated with people’s COVID-19 preventive measure-related knowledge, attitude, and practice. Studies regarding COVID-19 preventive measures in another geographical area might not mimic the same weight for these factors as it was done in the Rohingya refugee camps. According to the author’s knowledge, this is the second study that evaluates the KAP of Rohingya refugees in relation to COVID-19, allowing us to assess and compare our findings with those of a previous study conducted in Rohingya refugee camps and other refugee-related KAP studies. The results of this study indicate that the mean scores for knowledge and attitude were 9.93 (out of 14) and 7.55 respectively (out of 11). This reflects an increase in knowledge and attitude among Rohingya refugees compared to the results of previous research was done at the beginning of the pandemic [18], which indicated the mean score of 5.8 (out of 10) for knowledge and 2.2 (out of 5) for attitude. When it comes to quantifying the level of practice, our study found a mean score of 2.71 (out of 7) indicating that practice is not improved as much as knowledge and attitude. The practice level, however, is better than that found in the prior study; the mean practice score was 0.9. (Out of 5) [15]. The improvement of knowledge, attitude, and practice can be linked to the extensive risk communication and community engagement (RCCE) activities undertaken by the humanitarian agencies [19, 20]. The RCCE strategies of IOM and the health sector for COVID-19 include dissemination of information and engagement of community through household visits (with proper distancing), distribution of promotional materials, small group and courtyard meetings, and engagement of community leaders [23, 24]. Our study findings revealed that respondents possess adequate knowledge about COVID-19. In 2019, a survey was conducted on Rohingya people in the Cox’s Bazar camps addressing water, sanitation, and hygiene (WASH), and identical results were obtained in terms of total knowledge scores [26]. The study found that over half of the respondents were aware of COVID transmission which is similar to findings from Syrian refugee settlements [27]. In contrast, a previous study done at Rohingya camps found that the respondents were unable to answer questions about transmission [18].

Multivariable logistic regression analysis revealed a limited number of significant associations as measured by the adjusted odds ratio. In terms of knowledge level, respondents’ knowledge regarding COVID-19 infection prevention measures decreased with advancing age, and those who are elderly exhibited the lowest levels of knowledge which is similar to the earlier study conducted by Jubayer et el. [18]. Older persons are disproportionately affected by the emergencies and may have limited access to information [28]. In 2021, Lebrasseur et al. found that the COVID-19 pandemic had a major impact on vulnerable populations, notably older people, who often experience loneliness, age discrimination, and anxiety [28]. As a result, it is reasonable to assume that they have less knowledge than younger adults, which reflects in their degree of poor attitude and or practice, and the majority of elderly Rohingya respondents held a similar view of COVID-19 preventive measures, according to our findings. We found that participants generally had a positive attitude towards measures such as avoiding crowds, wearing masks in public, practicing physical distancing, and maintaining hand hygiene. However, it was also found that relatively few people actually put these attitudes into practice. Additionally, the hygiene practices of the Rohingya participants in the study seemed to differ from the findings of a previous study conducted in 2020 [22]. A study prior to the COVID-19 outbreak in the Rohingya refugee settlements found that more than 50% of Rohingya refugees had poor hygiene practices [26]. Our study found a strong correlation between good hygiene practices and family size. However, another study conducted during the early stages of the pandemic found that family size did not affect the hygiene practices of Rohingya people [29].

During the study period, we discovered that a significant number of participants over the age of 55, roughly two-thirds, received COVID-19 vaccine. This can be linked to the fact that to increase vaccine acceptance and uptake, Community Health Working Group (CHWG) implemented extensive risk communication and community engagement strategies along with evidence-based initiatives [30-31]. The study found that the participants did not take sufficient precautions, such as staying home and maintaining social distance, during the upsurge period. This lack of adherence to safety measures was similarly observed in another study conducted among Rohingya refugees, which found that over half of the participants did not maintain social distancing and frequently left their homes [22]. Conversely, the mothers of Syrian refugees exhibit a highly optimistic attitude, and they tend to follow social distancing protocols [27].

### Limitations and future directions

The design of this study was cross-sectional; hence, causal inferences may not always be made. The study found that many of the participants had a good level of knowledge and an average level of attitude, however, a significantly low level of practice of the COVID-19 preventive measures. While the study provides a numerical analysis, further qualitative investigations is required to understand the drivers and barriers of this situation. The quantitative findings of the study on different areas and level of knowledge, attitude and practice together with proposed qualitative investigation should be taken into consideration for updating the risk communication and community engagement strategy for COVID-19 and future similar outbreaks.

## Conclusions

The RCCE interventions improved the knowledge and attitudes of Rohingya refugees in Cox’s Bazar on COVID-19 preventative measures, but the results indicate that more effective interventions are required to improve knowledge and attitudes. It was also found that the score of practice, despite improving in comparison to the baseline, is significantly inadequate. There was a strong correlation between age group and knowledge level; older and younger age groups had less knowledge of COVID-19 prevention strategies. Further efforts should be made to determine the causes of the low level of practice, including the relationship between policies and tactics, the local context, resources, and social variables. Moreover, The problems of older adults’ low levels of knowledge should be investigated and addressed.

## Data Availability

All relevant data are within the manuscript and its Supporting Information files.

## Acknowledgements

The data for this article arose from the world’s largest refugee camps in Cox’s Bazar, Bangladesh. We would like to convey our heartfelt gratitude for the participant’s assistance.

## Funding

This research was supported by International Organization for Migration (IOM).

## Conflicting interests

The author(s) declared that they have no conflicts of interest in any steps of this research.

## Disclaimer

The views presented in the study are solely those of the authors and do not represent the official stance of the International Organization for Migration (IOM). This publication was issued without formal editing by IOM.

### Abbreviations

AWD: Acute Watery Diarrhoea
CHW: Community Health Worker
CI: Confidence Interval
CwC: Communication with Community
ERC: Ethical Review Committee
FDMN: Forcefully Displaced Myanmar Nationals
IOM: International Organization for Migration
IRB: Institutional Review Board
ISCG: Inter Sector Coordination Group (ISCG)
KAP: Knowledge, Attitude, and Practice
MERS-CoV: Middle East Respiratory Syndrome Coronavirus
NSU: North South University
RCCE: Risk Communication and Community Engagement
SARS-CoV-2: Severe Acute Respiratory Syndrome Coronavirus 2
WASH: Water, Sanitation and Hygiene
WG: Working Group
WHO: World Health Organization

## Notes

### Competing Interest Statement

The authors have declared no competing interest.

### Funding Statement

This work was not specifically funded by the author(s) but was supported by the International Organization for Migration (IOM).

### Author Declarations

The Institutional Review Board (IRB)/Ethical Review Committee (ERC) of North South University, Bangladesh, authorized the protocol for this study (2021/OR-NSU/IRB/0401).

